# Anaesthesia delivery systems in low and lower-middle-income Asian countries: a scoping review of capacity and effectiveness

**DOI:** 10.1101/2023.05.03.23289468

**Authors:** Sumbal Shahbaz, Natasha Howard

## Abstract

**Background:** Literature on anaesthesia systems in low and lower middle-income countries is limited, focused on the Africa region, and provides minimal data on anaesthesia or associated disciplines within intensive care, pain management and emergency medicine. We thus conducted a review of primary and secondary research literature on low and lower middle-income countries in the Asia region from 2000-2021, to clarify existing knowledge, important gaps, and possible subsequent steps.

**Methods:** We applied Arksey and O"Malley"s scoping literature review method to search, screen, extract, and synthesise data under three themes: (i) availability and type of anaesthesia workforce; (ii) anaesthesia system infrastructure, equipment, and supplies; and (iii) effectiveness of anaesthesia provision.

**Results:** We included 25 eligible sources of 603 identified. Only ten (40%) were published in the last 5 years and Asian lower-income countries were primarily represented in 15 multi-country sources. Fifteen (60%) sources used quantitative methods and provided limited information on data collection, e.g. sampling criteria or geographic areas included. No sources included countrywide data, despite anaesthesia delivery and resources differing significantly sub-nationally (e.g., central versus rural/remote, or insecure areas). Data on anaesthesiology delivery were limited, with findings including insufficiencies in workforce, supplies, training and skills-building of anaesthesia personnel, along with the lack of consistent strategies for overcoming maldistribution of resources and improving anaesthesia delivery systems in the region.

**Conclusions:** This review, a first attempt to synthesise existing data on anaesthesia delivery systems in low and lower-middle-income Asian countries, shows the anaesthesia literature is still limited. Findings highlight the urgent need for additional research and collaboration nationally and regionally to strengthen anaesthesia delivery and surgical facilities in resource-constrained settings.

## INTRODUCTION

Anaesthesia provision that is timely and of good quality can significantly reduce surgical mortality and morbidity, yet almost 90% of populations in low and middle-income countries (LMICs) have difficulties accessing surgical care (1) due to insufficient availability of quality anaesthesia, making it a major limitation globally in achieving comprehensive surgical care needs. This can cause delayed or no surgical treatment for many common and treatable conditions, such as appendicitis or obstructed labour, resulting in higher mortality rates (2). Additionally, many people in LMICs receive anaesthesia from untrained or unskilled anaesthesia providers, resulting in higher mortality rates than in high-income countries (3).

It is challenging to follow improvements in accessing quality anaesthesia services over time in LMICs (4), especially due to the lack of research in the Asia region, migration of skilled personnel, maldistribution of resources, and protracted or unanticipated conflict and occupation (e.g. Afghanistan, Syria, Palestinian territories). International anaesthesiology research and funding for LMICs is focused primarily and understandably on resource-depleted settings in the Africa region, with most documentation of anaesthesia capacity and effectiveness over time conducted in African countries. Despite less documentation, anaesthesia provision needs also exist in the Asia region, requiring efforts to improve numbers of trained anaesthesia personnel, equipment, and medication (5). Many lower-income Asian countries have insufficiently resourced systems, with as few as 9 anaesthetists for a population of 32 million in Afghanistan (6).

We contend that existing knowledge on anaesthesia provision needs in lower-income Asian countries require synthesis to highlight potential knowledge and practice gaps that could be addressed through additional research and funding. Given the insufficient international prioritisation of research data, we conducted a scoping literature review of the capacity and effectiveness of anaesthesia delivery systems in lower-income Asian countries, examining infrastructure, workforce, and services. This review can provide a starting point for advancement in this important yet neglected area of medicine in LMICs.

## METHODS

### Study design and definitions

We conducted a scoping literature review using Arksey and O"Malley"s method and later refinements (7-11). We chose a scoping method given the breadth of our research question and anticipated heterogeneity of the literature (9), because it can legitimately be conducted by a single investigator, and because it does not restrict data through formal quality assessment (10,11).

Table 1 shows our definitions. We used Tranquilli and Thurmon"s 2013 anaesthesia definition, as it broadly defines the term and, although anaesthesia is no longer limited to surgical procedures, it is still largely limited to intraoperative procedures in LMICs. Similarly, our chosen definition for anaesthesia delivery system refers to its intraoperative use (12).

**Table 1.** Research definitions.

|  |  |
| --- | --- |
| Anaesthesia | Anaesthesia is categorized as insensitivity to pain, particularly by induction of synthetically-prepared gases or inoculation of drugs prior to surgical procedures (13). |
| Anaesthesia delivery system | The anaesthesia delivery system comprises the anaesthesia provider, anaesthesia machine, anaesthesia vaporizers, ventilator, breathing circuit, and waste gas scavenging system (12). |
| Health system | Consists of all organizations, people and actions whose primary intent is to promote, restore or maintain health, including efforts to influence determinants of health as well as more direct health-improving activities (WHO, 2007) |
| Lower-income Asian countries | Include low-income countries, with a per capita gross national income (GNI) of less than US\$1045 annually, and lower-middle-income countries with a GNI per capita of US\$1046-4096 annually (OECD, 2022). In the Asia region, these are Afghanistan, Bangladesh, Bhutan, Cambodia, India, Indonesia, Iran, Kyrgyzstan, Laos, Mongolia, Myanmar, Nepal, North Korea (DPRK), Pakistan, Palestine, Papua New Guinea (PNG), Philippines, Sri Lanka, Syria, Tajikistan, Timor-Leste, Uzbekistan, Vanuatu, Viet Nam, and Yemen (World Bank, 2022). |

We chose the standard WHO health system definition for its familiarity. For lower-income Asian countries, we chose 24 identified by the Organisation for Economic Co-operation and Development (OECD) list for 2022-2023 as low-income or lower-middle-income (i.e. below US$4045 annual GNI) among the 48 Asian countries recognised by the United Nations.

### Research question

Our research question was: „What is the scope (i.e. extent, nature, distribution) and main capacity and effectiveness findings of the existing literature on anaesthesia delivery within healthcare in Asian low and lower-middle-income countries?”

### Identifying relevant sources

To ensure breadth and comprehensiveness, we searched five electronic databases systematically (i.e. EMBASE, CINAHL, Medline, Scopus, Web of Science), using the terms and related terminology for „anaesthesia” AND „delivery system” AND „LMICs” AND „Asia” adapted to the subject headings for each database. Table 2 provides an example in Medline.

**Table 2.** Search syntax and keywords for Medline.

| Key word | Medline |
| --- | --- |
| Anaesthesia | 1. "Anesthesiology"[Mesh]<br>2. "Anesthesia"[Mesh]<br>3. anesthesiology OR anaesthesiology OR<br>4. anesthesia OR anaesthesia<br>5. 1 OR 2 OR 3 OR 4 |
| Health system | 6. Delivery of Health Care / methods<br>7. Delivery of Health Care / standards*<br>8. Public Health / methods OR Public Health / standards<br>9. Quality of Health Care / standards*<br>10. 6 OR 7 OR 8 OR 9 |
| Asian LMICs | 11. Low- and Middle-Income Countries<br>12. Developing Countr* OR Developing Nation* OR Less-Developed Nation*<br>13. Least Developed Countr* OR Less-Developed Countr* OR Under-Developed Nation* OR Under-Developed Countr* OR<br>14. Third-World Nation* OR Third-World Countr* OR<br>15. 11 or 12 or 13 or 14<br>16. Afghanistan OR Cambodia OR Azerbaijan OR Bangladesh OR Bhutan OR India OR Indonesia OR Iran OR Democratic people's republic of Korea OR DPRK OR Mongolia OR Myanmar OR Nepal OR Pakistan OR Palestine OR West Bank and Gaza OR Papua New Guinea OR Kyrgyzstan OR Lao* OR Philippines OR Sri Lanka OR Syria OR Tajikistan OR Timor-Leste OR East Timor OR Uzbekistan OR Viet Nam OR Vietnam OR Yemen<br>17. 5 AND 10 AND 15 AND 16 |

### Selecting sources

Table 3 provides eligibility criteria, determined via an iterative process. Context was restricted to Asian LMICs to help inform anaesthesiology in the region. Topic was restricted to anaesthesia delivery system defined in Table 1. Outcomes were restricted to capacity and effectiveness measures. Source type was restricted to primary and secondary research literature. Time-period was restricted to 2000 and after, as before this anaesthesia practices, equipment, and medications were sufficiently different to affect research findings. All languages, study designs, and participants were considered.

**Table 3.**
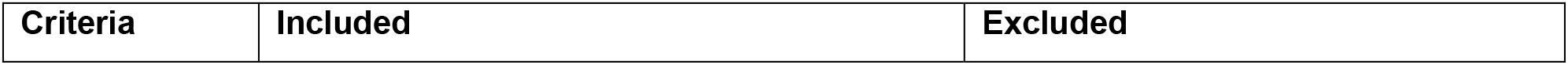
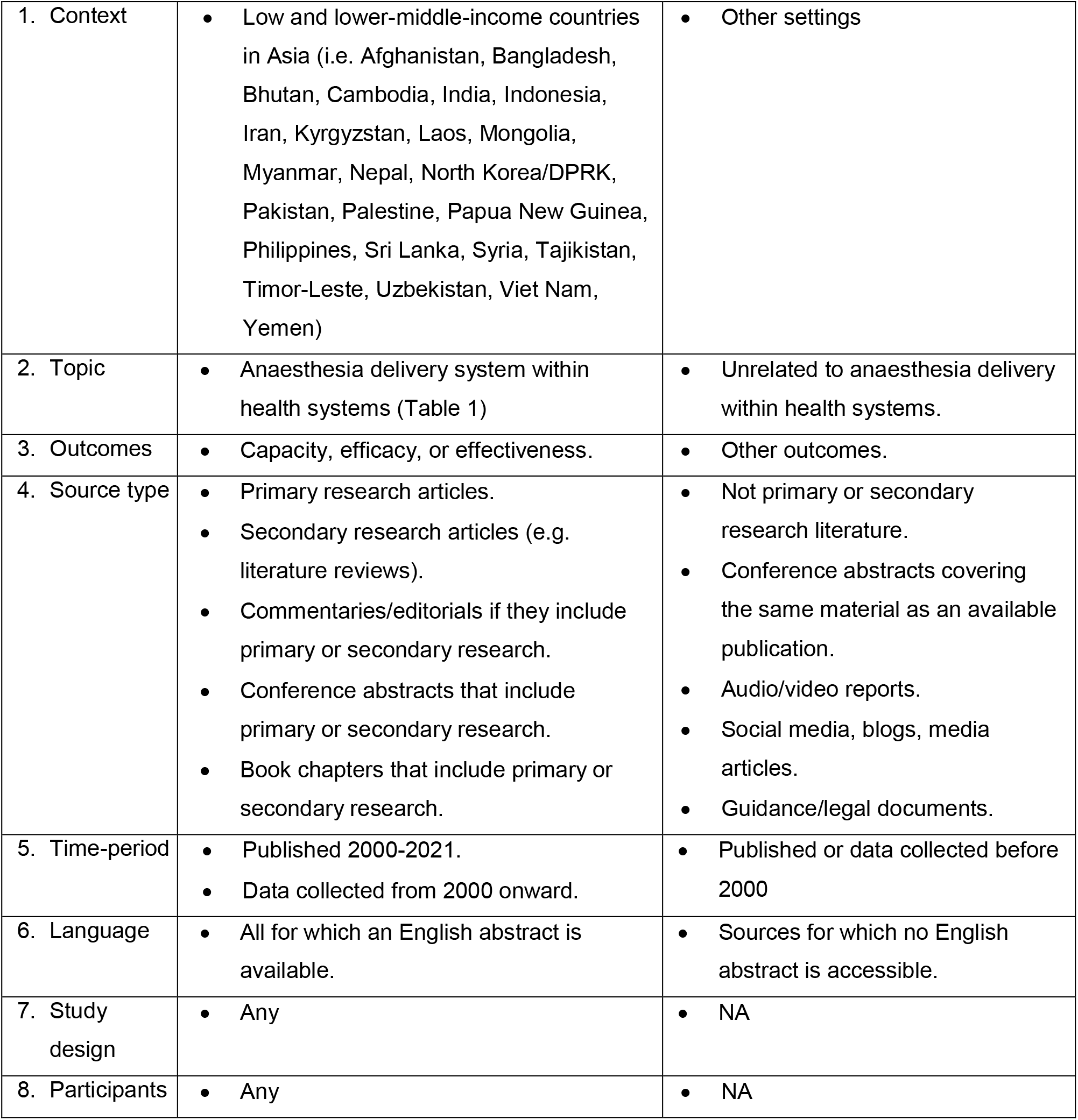
Eligibility criteria.

First, we downloaded all sources identified across the five databases into EndNote reference manager and deleted all duplicates. Second, we screened titles and abstracts against eligibility criteria and eliminated obviously ineligible sources using Rayyan software. Third, we screened full texts against eligibility criteria and eliminated ineligible sources. Finally, we screened reference lists of included sources to identify any additional eligible sources. This provided our total number of sources included.

### Extracting (charting) data

We extracted data from eligible sources to an Excel sheet using the following iterative headings: (i) source identifiers, i.e., publication year, lead author, source type (e.g., article, conference abstract, report), language; (ii) source characteristics, i.e., country/ies, study design, participant characteristics, methods; (iii) findings, i.e., capacity (workforce, infrastructure), efficacy, and effectiveness.

### Synthesising and reporting results

First, we summarised the extent (i.e. numbers, publication year, type - article, conference abstract, book, report), distribution (i.e. publication language, countries included), and nature (i.e. study design, participants, outcomes) of sources. Second, we synthesised findings thematically, guided by Braun & Clarke"s approach, under three deductive themes: (i) availability and type of anaesthesia workforce; (ii) anaesthesia infrastructure, equipment, and supplies, (iii) effectiveness of anaesthesia provision (15).

## RESULTS

### Scope of the literature

#### Extent

We included 25 eligible sources of 603 identified in databases and reference lists. Most were from EMBASE (195) and Medline (185), 209 and 84 records were removed by title/abstract and full-text screening respectively, while 6 were added from purposively searching reference lists of included sources.

#### Distribution

All sources were published in English. Data from all 24 lower-income Asian countries (100%) were included. Ten (40%) were conducted in individual countries while 15 (60%) were conducted in LMICs globally and included 24 lower-income Asian countries. Multi-country sources included data from 5 to 24 countries. Figure 3 shows, Afghanistan was most represented (1 single, 7 multi-country sources); followed by Bangladesh (2 single, 4 multi-country) and India (3 single, 3 multi-country); Pakistan and Sri Lanka (1 single and 4 multi-country each); Viet Nam (4 multi-country); Bhutan, Cambodia, Indonesia, Iran, and Nepal (3 multi-country each); Papua New Guinea (1 single, 2 multi-country); while Syria was in one single and one multiple-country source; and Myanmar, North Korea, and Philippines were in 2 multi-country sources. Kyrgyzstan, Laos, Mongolia, Palestine, Tajikistan, Timor-Leste, Yemen, and Uzbekistan were represented in only one multi-country sources. India had the most single-country sources, with 3 conducted in separate states.

**Figure 1.**
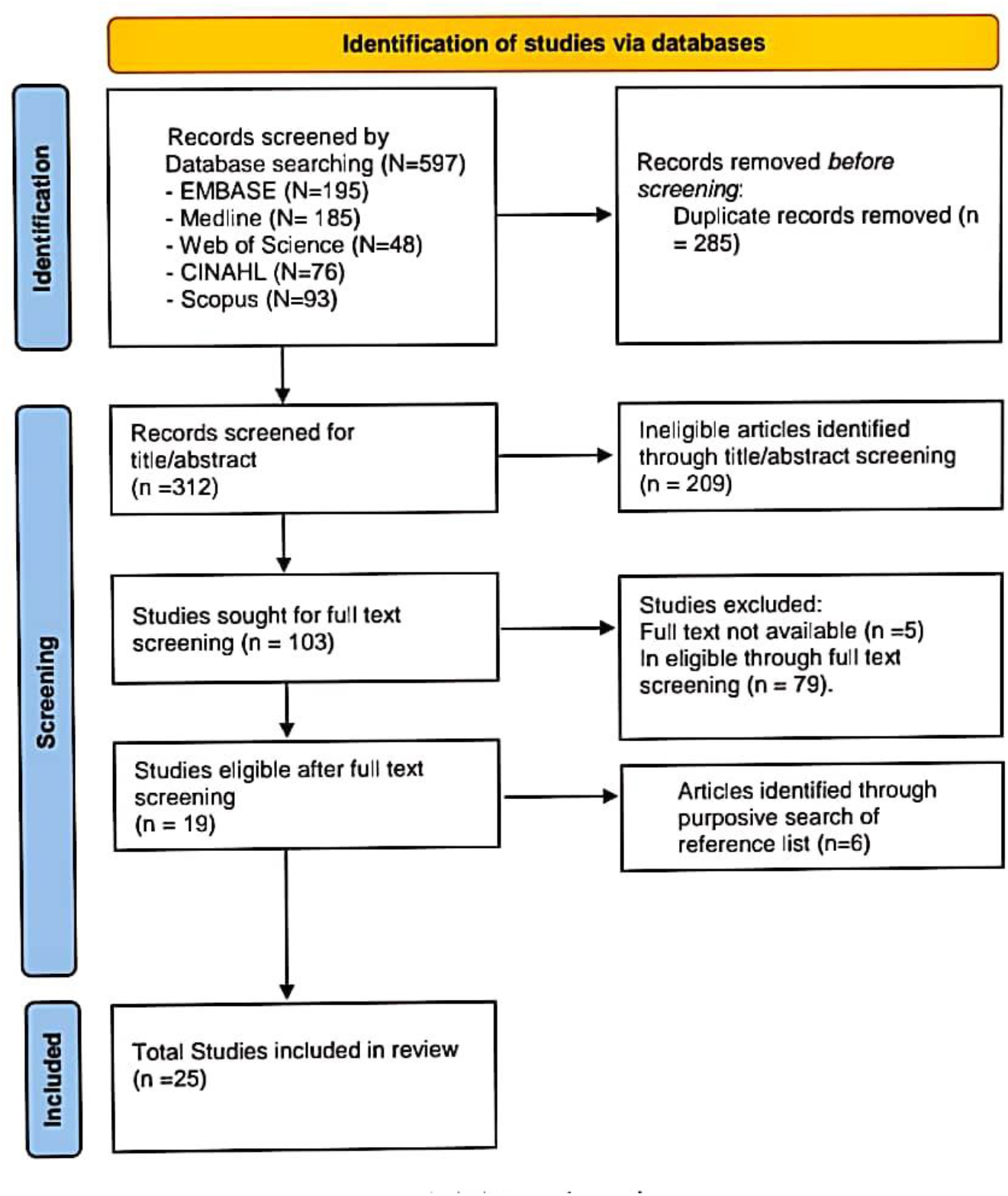
PRISMA flow diagram. No sources were published prior to 2006, or in 2008 and 2011, while the maximum number per year never exceeded 3 (e.g. in 2010, 2015, 2017, and 2020 respectively). Overall, no clear trend emerged.

**Figure 2.**
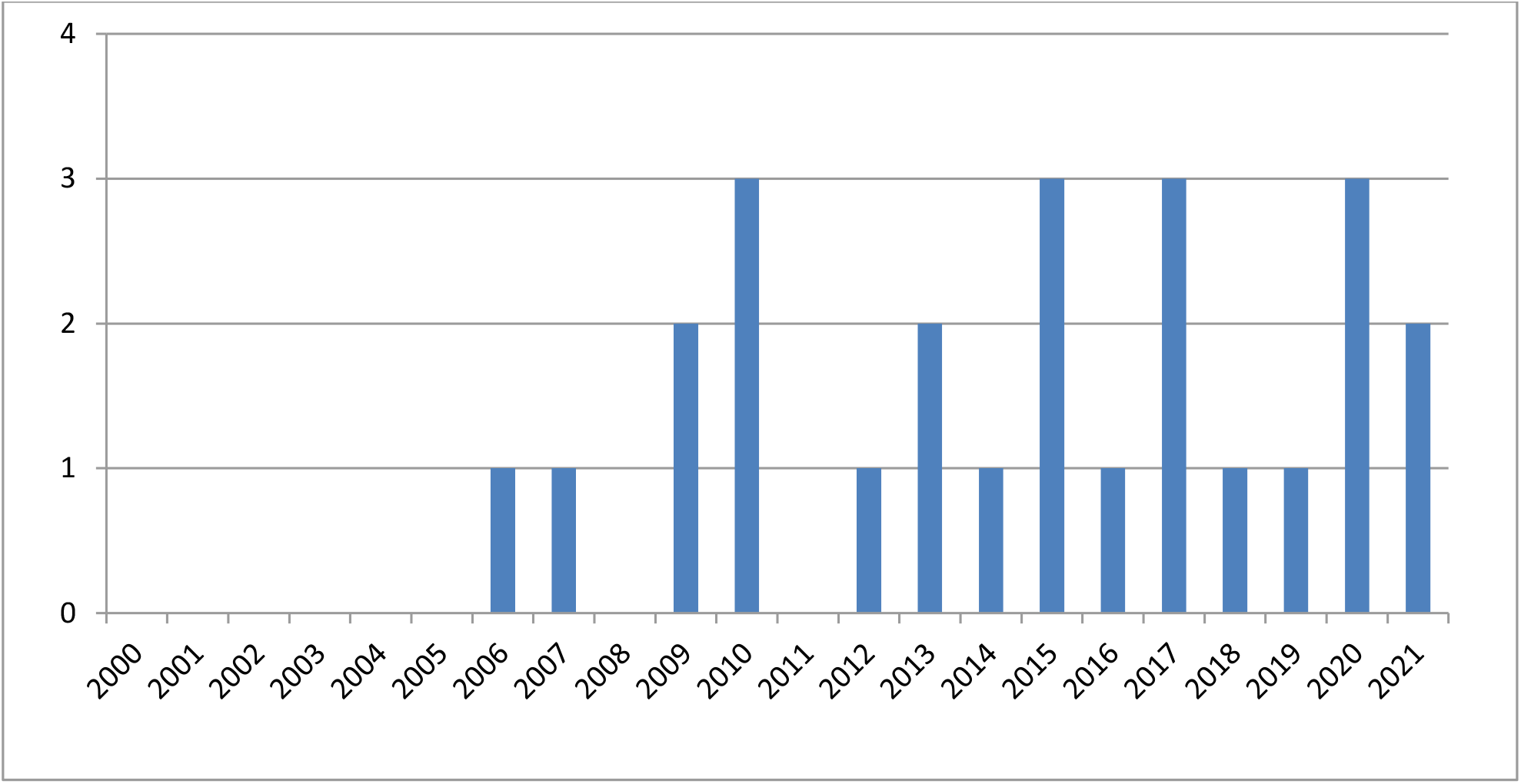
Publication numbers by year. Most sources (17/25; 68%) were research articles, while 6 (24%) were review articles and 2 (8%) were technical reports.

**Figure 3.**
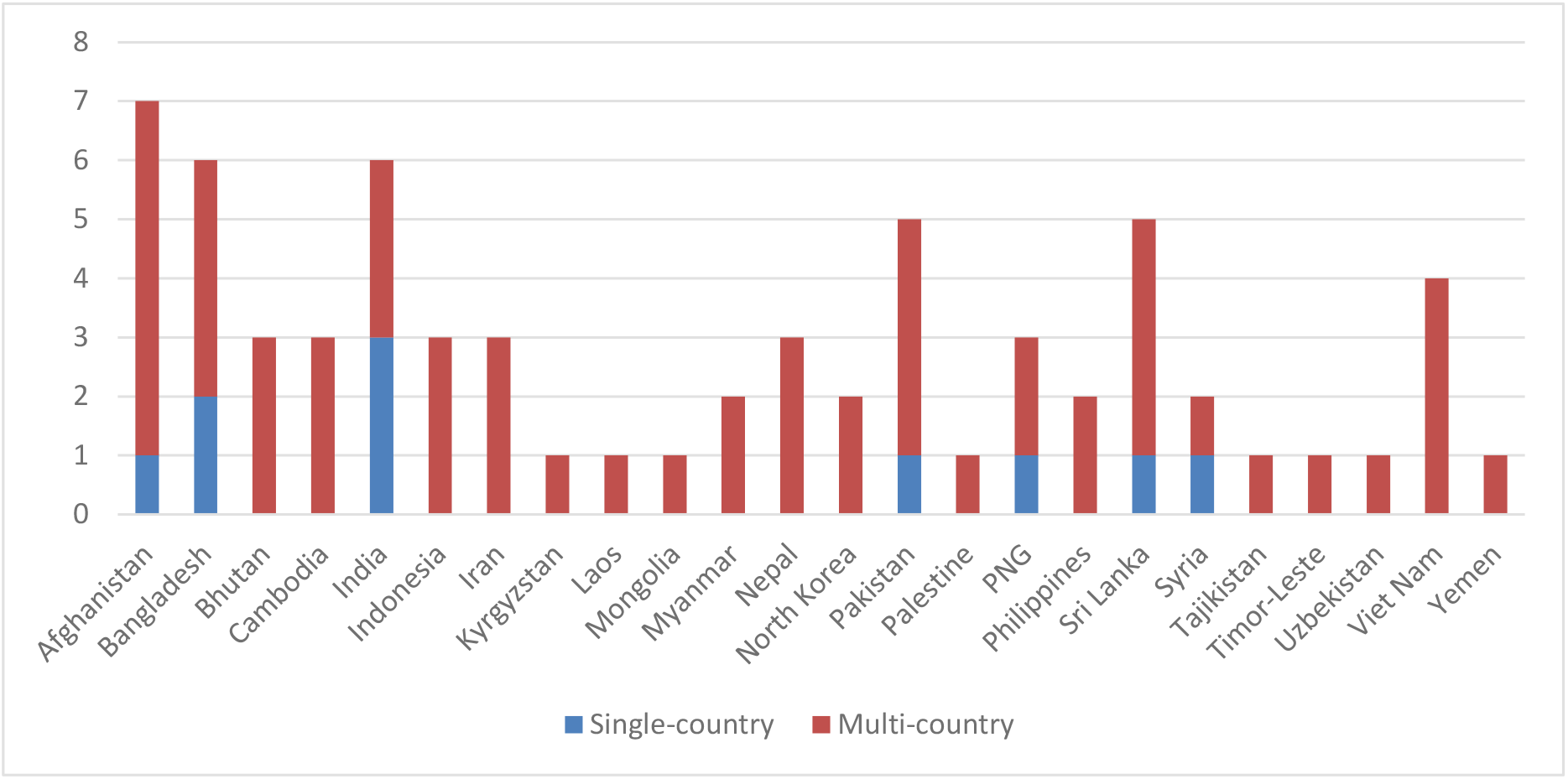
Publications numbers by country.

#### Nature

Study methods for 15 (60%) articles were quantitative, primarily cross-sectional (multicentre or observational) surveys. Mostly used WHO or World Federation of Societies of Anaesthesiologists (WFSA) approved survey tools and participants were primarily anaesthesia personnel from health facilities who enumerated available resources and described practices. Two (8%) sources primarily used qualitative methods, including semi-structured interviews and observations. A few sources detailed their sampling strategy, while most provided minimal explanation. Random and purposive samplings were the preferred sampling methods.

### Synthesised findings

Table 4 shows coverage of our three deductive themes by sources. Most (21) included more than one theme, and 13 included all three, though depth and rigour varied considerably.

**Table 4.**
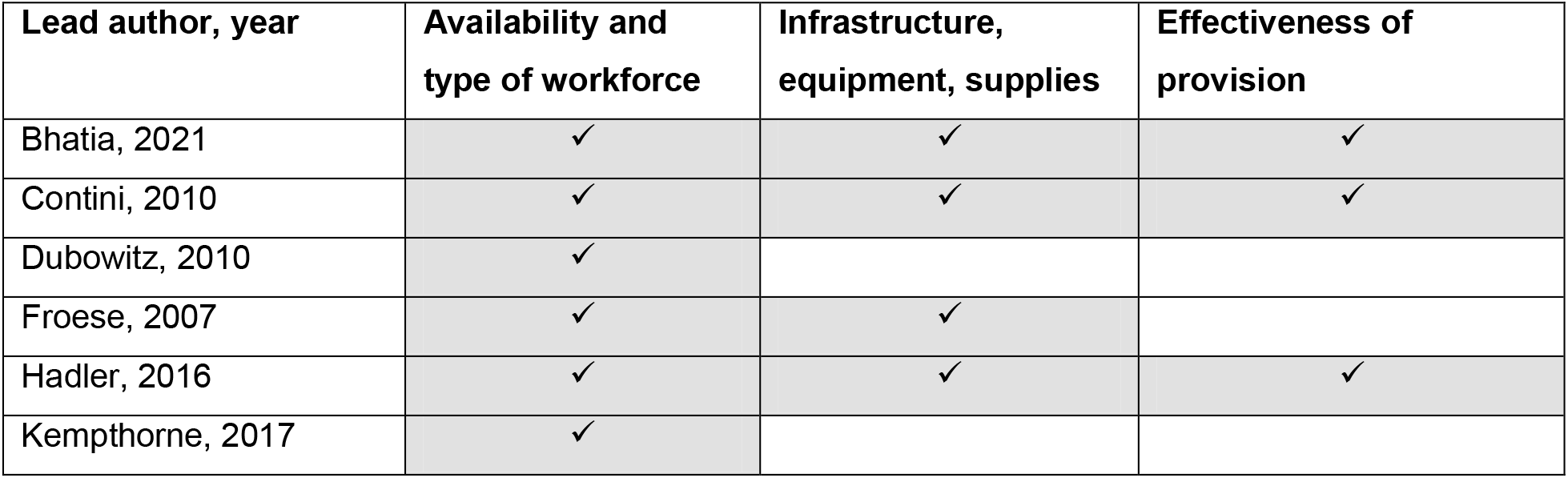
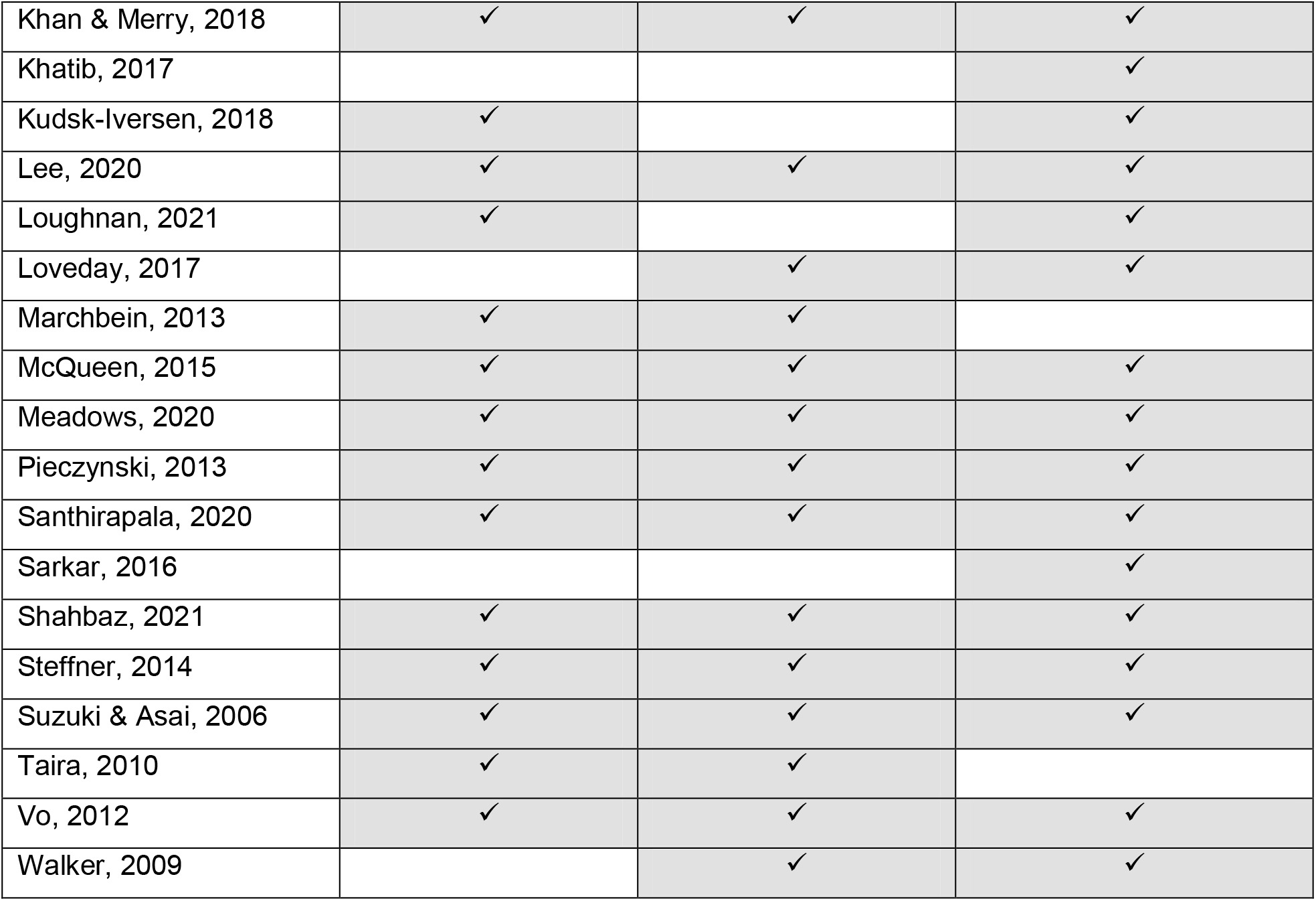
Coverage of themes by source.

### Availability and type of workforce

Twenty sources (80%) included data on this theme, all indicating a lack of sufficient trained anaesthesia personnel. Anaesthesia workforce shortages were acknowledged repeatedly as a key obstacle in achieving safe surgical care. Dubowitz and colleagues reported anaesthesia workforce numbers as low as 0.07 per 100,000 population in Yemen, including physician and non-physician anaesthetists (16). Similarly, Steffner et al noted the absence of even mid-level anaesthesia providers (i.e. dedicated anaesthesia nurses or technicians) in many hospitals they surveyed, leading to anaesthesia provision by physicians and non-physicians without any formal anaesthesia training, and increased perioperative death rates (17). Contini et al found only 5 of 17 health facilities assessed in Afghanistan had trained physician-anaesthetists, while a few had nurse anaesthetists, but most had no anaesthesia personnel (18). In 22 LMICs reviewed by Hadler et al, only 56% of hospitals had capacity to perform general anaesthesia due to a general lack of trained anaesthetists (19).

Vo and others found anaesthesia provider numbers increased according to hospital bed numbers rather than need or population served, so a 300-bed hospital could have almost 4 anaesthetists compared to 100-bed or smaller hospitals with less than 1 fulltime or no anaesthetist to provide services to a larger population (20). Bhatia et al reported 1 anaesthetist in a Haryana sub-district hospital for a population of 500,000, which not only showed the lack of anaesthesia workforce but also delayed surgeries due to the lack of appropriate anaesthesia personnel in parts of India (21). Likewise, Loveday et al described 952 anaesthetists for a population of 163.05 million in Bangladesh (i.e. averaging 0.58 anaesthetists per 100,000 people), a minor increase from 0.52 per 100,000 in 2012 (22). In most LMICs, non-physician anaesthetists were not routinely trained to help support the system. However, Papua New Guinea, which has only 0.25 physician-anaesthetists per 100,000 populations, trained non-physician anaesthetists to meet 90% of total anaesthesia demand (23). Meara and others thus recommend task sharing with non-physician anaesthetists, who are cheaper and quicker to train to fill the gaps in LMICs until minimum essential standards have been attained (2, 24, 25).

No sources covered any country fully and consultants in most countries are concentrated in city centres (26). Thus, the situation is likely worse in rural, hard-to-reach, and insecure or conflict-affected areas (6). Moreover, many skilled anaesthetists reportedly left the speciality or their country due to high workloads, burnout, insecurity, or feeling undervalued/under-remunerated (Kudsk-Iversen, et al. 2018).

### Infrastructure, equipment, and supplies

Eighteen sources (72%) included data on this theme, with none reporting 100% availability of uninterrupted water, oxygen, or electricity in the health facilities examined (18, 27).

Access to fully-functional anaesthesia equipment was limited to half of surveyed facilities in most countries (17, 20). Likewise, Walker et al found availability of pulse oximeters was limited to approximately half of health facilities in Viet Nam and the Philippines (28). A volunteer anaesthetist in India mentioned his concerns about losing his patient due to the lack of basic monitoring equipment (29).

Sources found majority of facilities relying on Ketamine due to constrained resources or use regional anaesthesia only, owing to the lack of emergency intubation equipment (18, 19).

Moreover, Contini et al found paediatric intubation sets were not available in half of health facilities examined in Afghanistan (18). Availability of blood banks and invasive monitoring was restricted to tertiary care facilities in Pakistan and other resource-depleted countries (24, 26). Face masks, bags, ECG monitoring and medication were reported as absent in most health facilities studied in Bangladesh (22). Infrastructure, supplies, and medications availability were reportedly worst in primary and secondary hospitals in almost all sources with only teaching and tertiary hospitals having necessary anaesthesia supplies available, though still often in limited quantities (30).

### Effectiveness of anaesthesia provision and future interventions

Nineteen sources (76%) included data on this theme. Despite insufficiencies in workforce and supplies described, consistent strategies for improving the anaesthesia delivery system in lower-income countries in the region appeared to be lacking. This lack of strategic direction was particularly noticeable in the absence of anaesthesia monitoring and evaluation data collected or analysed, insufficient training and skills building of anaesthesia personnel, frequent maldistribution of anaesthesia resources, and the added health system burden of armed conflict and insecurity.

Contini and others found subnational difference in performance data and outcomes in Afghanistan and other countries, showing the urgent need for nationwide anaesthesia data collection and analysis in each country (18, 20). Steffner et al noted that any improvement in the anaesthesia system is impossible until comprehensive data analysis on clinical outcomes, cost-effectiveness, mortality, and morbidity, while reporting the absence of appropriate indicators on anaesthesia access and outcomes in countries studied (17). Some sources suggested the perioperative mortality rate could be the WHO-recommended health indicator to monitor and compare perioperative infrastructure across health systems (25).

The presence of anaesthesia personnel did not automatically determine the provision of quality anaesthesia services, as theoretical knowledge and skills could be outdated or insufficient (25), leadership may be lacking, and essential equipment and supplies may be unavailable. Some sources focused on the importance of improving anaesthesia training along with capacity improvements (19). Nurses/technicians delivered most anaesthesia in non-central areas in the countries included, or junior doctors with variable training, who also trained other personnel in anaesthesia delivery despite their own limited skills. Thus, if anaesthesia providers training and capacity are considered, the dearth of anaesthesia effectiveness is even greater. Combining this limited knowledge with limited resources could contribute greatly towards perioperative and intraoperative mortality rates. One source described the positive aspects of involving anaesthesia volunteers from high-income settings in building capacity in LMICs, as not only improving patient care but also training LMIC anaesthesia professionals to continue the same standard of care in their own countries (29).

One of the most important aspects mentioned in sources was the mal-distribution of anaesthesia resources that made remote and secondary health facilities unsafe for anaesthesia provision (22, 27, 30). Several sources reported better facilities and concentrations of qualified anaesthetists in teaching and tertiary level hospitals, while health facilities in remote areas were left without meaningful guidance or support (22, 27, 30).

Moreover, many Asian countries did not allow employment of non-physician anaesthetists, which not only promoted unsafe practices but also burnout among the limited available physician-anaesthetists. With growing focus on the global surgery agenda, anaesthesia could likely become a rate-limiting step to increasing surgery capacity in many LMICs in Asia (22).

Contini and others described how decades of war in some Asian LMICs further degraded the anaesthesia delivery system, with either no data available on surgery and anaesthesia or data going unreported or being misplaced or destroyed (18). Marchbein et al also reported that workable pre-conflict anaesthesia delivery systems were often destroyed or evacuated during conflict. For example, security concerns in Syria meant health-workers were unwilling to share data as this could make them targets for treating opposition fighters (31).

## DISCUSSION

### Key findings

This review is the first to our knowledge to synthesises the scope and main findings of the literature on capacity and effectiveness of anaesthesia delivery systems in lower-income Asian countries. Eligible sources were limited, as most anaesthesia delivery literature only discussed clinical aspects, but highlighted important delivery weaknesses that contributed to preventing the region from achieving international criteria for minimum essential anaesthesia staffing, equipment, and medication. The relatively limited literature indicates the need for further research on this topic in Asia. However, most sources from different countries discussed common trends and issues, which enabled thematic synthesis. The review thus provides a starting point for future research and analysis on anaesthesia delivery systems in lower-income economies in the region.

Availability of oxygen, water, and electricity are minimum standards for facilities providing surgery and anaesthesia, but no single country included in our review was providing this 100% of the time in all facilities (32). This lack of infrastructure was in line with several studies in the Africa region, showing 50-75% of African hospitals assessed were without basic facilities such as pulse oximeters or monitors (33-35). In Asian LMICs, 50-75% of hospitals studied had these facilities except those experiencing on-going conflict(26). Most teaching and tertiary-level hospitals in our review met WFSA mandatory standards for safe practices including availability of opioid analgesics (36), while secondary and primary health facilities generally lacked these as also noted for LMICs in South America and Africa (34, 37-39). As found previously in LMICs, ketamine was extensively used in many countries (40).

This emphasises the importance of ketamine in LMICs until international criteria of minimum essential equipment and medication have been met (41).

WFSA recommends a minimum of 5 physician-anaesthetists per 100,000 population (6). Unfortunately, even after including non-physician anaesthetists, none of the countries in this review achieved this target. In most of these countries, non-physician anaesthetists are not allowed to practice or given any formal training to overcome this gap as compared to many African countries in which anaesthesia officers and nurse-anaesthetists work effectively to bridge the gap in qualified personnel (42-44). Poor employment conditions, security, burnout, and limited professional acknowledgment are noteworthy obstacles to recruitment and retention of the anaesthesia workforce in Asian LMICs (45). The stress, high-workloads, insufficient remuneration, and security issues associated with anaesthesiology reduces the number of medical students joining this speciality, while migration of skilled anaesthesia professionals to high-income countries replicates the health-worker „brain-drain” found in many health specialisations (46, 47).

Key implications for policymakers and, practitioners are the urgent need to achieve minimum anaesthesia standards for infrastructure and staff in the region. First steps in improving these anaesthesia systems would be strengthening adherence to WHO guidance on data recording and reporting and assessing and standardising capacity in numbers, training, and experience of physician and non-physician anaesthetists(48). It should thus be noted that several eligible countries, i.e. Kyrgyzstan, Mongolia, Palestine, Tajikistan, Uzbekistan were not included in any sources regarding anaesthesia capacity (i.e. equipment and medication) or effectiveness, which could threaten achievement of the global plan of surgical and anaesthetic safety (49). Thus, a key role for researchers would be to support efforts to fill data gaps, particularly for these countries.

### Limitations

Several limitations should be considered. First, the literature was heterogeneous, with substantial differences in methodologies, data collection tools, and study samples. Second, no source represented an entire country, despite the acknowledgement of significant subnational differences in anaesthesia resources and capacities, particularly between capital and rural or remote regions. Third, study quality assessment was not conducted as it is not required for scoping reviews and could have further reduced the number of eligible sources. Fourth, individual country-based data from only 7 countries were available, indicating further data collection is required in all countries to better understand the situation in the region.

However, several patterns were clear despite data gaps, including the lack of electricity, oxygen, and pulse oximeters, insufficient quantity and quality of anaesthesia staff, and challenging work environments. Finally, all assessments were conducted and reported by different research teams, which may have resulted in discrepancies in reporting. However, most tools were validated for multi-country use by WFSA or WHO.

### Conclusions

This is a first attempt to synthesise existing research data on anaesthesia health/delivery systems in Asian Lower income countries, which have often been overlooked due to more extreme health disparities in other regions. However, this review highlighted the urgent need for additional research and improved anaesthesia service quality in this region. Operational challenges in accessing remote and disputed or conflict-affected settings must be addressed, requiring collaboration among national and international organisations interested to improve anaesthesia and surgical facilities globally and in the region. Governments and partner organisations must budget some additional resources for improved data collection, training, and provisioning within anaesthesia systems if we are to prevent anaesthesia from being a rate-limiting step in surgery provision.

## Data Availability

All data were obtained from academic literature available in electronic databases.

## DECLARATIONS

### Author contributions

SS and NH conceived the study. SS conducted the review with supervision by NH. SS drafted the manuscript. NH provided critical revisions. All authors approved the version for submission.

